# Quantifying mutant huntingtin protein in human cerebrospinal fluid to support the development of huntingtin-lowering therapies

**DOI:** 10.1101/2022.07.22.22277927

**Authors:** Stephanie Vauleon, Katharina Schutz, Benoit Massonnet, Nanda Gruben, Marianne Manchester, Alessandra Buehler, Eginhard Schick, Lauren Boak, David J Hawellek

**Author notes:** **Corresponding author:** David J Hawellek, Roche, Pharmaceutical Research and Early Development, Roche Innovation Center Basel, Basel, Switzerland.

## Abstract

**Background:** Huntington’s disease (HD) is caused by a cytosine adenine guanine-repeat expansion in the huntingtin gene. This results in the production of toxic mutant huntingtin protein (mHTT), which has an elongated polyglutamine (polyQ) stretch near the protein’s N-terminal end. The pharmacological lowering of mHTT expression in the brain targets the underlying driver of HD and is one of the principal therapeutic strategies being pursued to slow or stop disease progression. This report describes the characterisation and validation of an assay designed to quantify mHTT in the cerebrospinal fluid of individuals with HD, for use in registrational clinical trials.

**Methods:** The assay was optimised, and its performance was characterised with recombinant huntingtin protein (HTT) varying in overall and polyQ-repeat length.

**Results:** The assay was successfully validated by two independent laboratories in regulated bioanalytical environments and showed a steep signal increase as the polyQ stretch of recombinant HTTs pivoted from wild-type to mutant protein forms. Linear mixed effects modelling confirmed highly parallel dose-response curves for HTTs, with only a minor impact of individual slopes of the dose-response for different HTTs (typically <5% of the overall slope). This implies an equivalent quantitative signal behaviour for HTTs with differing polyQ-repeat lengths.

**Conclusion:** The reported method may be a reliable biomarker tool with relevance across the spectrum of HD mutations, which can facilitate the clinical development of HTT-lowering therapies in HD.

**Trial registration:** Not applicable.

**Funding:** F. Hoffmann-La Roche Ltd.

## Introduction

Huntington’s disease (HD) is a rare, genetic neurodegenerative disease that is characterised by a triad of cognitive, behavioural and motor symptoms (1,2). Initial changes in brain pathophysiology underlie the early stage of HD. As the disease progresses, cognitive and motor symptoms become clinically detectable, followed by continued decline in body function (3).

HD is caused by a cytosine adenine guanine (CAG)-repeat expansion in the huntingtin gene (*HTT*), which is a direct determinant of potential or confirmed HD onset. A CAG-repeat length of ≥40 causes HD, while a CAG-repeat length of ≤26 does not. The middle ranges of 27–36 and 36–39 CAG repeats are known as ‘higher normal’ and ‘reduced penetrance’ respectively; the former will not cause HD but may be passed to subsequent generations, and the latter may or may not cause HD in the individual’s lifetime (1,4-6).

The CAG-repeat expansion results in the production of toxic mutant huntingtin protein (mHTT), which has an elongated polyglutamine (polyQ) stretch near the protein’s N-terminal end (1,4,7). Levels of mHTT in cerebrospinal fluid (CSF) correlate with disease stage, symptom severity and markers of neuronal damage in people with HD (8,9). Lowering mHTT production, via the degradation of *HTT* mRNA for example, targets the underlying driver of HD and interferes with the direct causal pathway of the disease. Consequently, mHTT is a key biomarker of HD as it has a direct causal involvement in the pathophysiology of the disease. This renders it a direct target for pharmacological interventions (4).

Huntingtin protein (HTT) has an extremely low abundance in the CSF (10), making ultra-sensitive platforms the most suitable method for detection. A novel, ultra-sensitive single molecule counting (SMC) mHTT immunoassay on the Erenna® platform was shown by Wild, et al (2015) to quantify CSF mHTT in association with proximity to disease onset and reductions in cognitive and motor function (8). Subsequently, evaluation of the assay in a research-grade environment suggests it may support the application of mHTT quantification as a biomarker in HD clinical trials for HTT-lowering therapies (11,12). A research-grade version of the assay has been used to analyse HD samples from the Phase I/IIa study of the antisense-oligonucleotide (ASO) tominersen (NCT02519036) (12).

The ligand binding assay uses capture antibody 2B7 which binds to both mHTT and wild-type HTT (wtHTT), and detection antibody MW1 which binds to the extended polyQ stretch of mHTT. Although MW1 is used in many available assays, the specificity of MW1 for mHTT remains relatively unclear. MW1 may have differential binding properties depending on the sub-cellular location of mHTT (13) as well as the exact number of CAG repeats in HTT (11), suggesting that not all HTT species are equally detected by MW1. The current study extensively characterised the assay response via experiments with recombinant HTT and patient CSF across a wide range of different polyQ lengths. Furthermore, the validations performed in separate laboratories demonstrate the inter-laboratory performance and replicability of the assay. In this paper, we report the optimisation and adaptation of the previous assay procedure on the SMCxPRO™ platform, along with two independent method validations according to international regulatory guidelines.

## Results

### Assay optimisation

Capture antibody 2B7 was originally labelled with biotin and detection antibody MW1 was labelled with the fluorescent dye Alexa Fluor® 647 according to SMCxPRO™ labelling kit manufacturer instructions, leading to high variability in assay performance. Optimisation of purification and labelling protocols together with thorough analytical characterisation of starting materials and end products improved performance and batch consistency of the assay reagents (**Supplemental Table 1**). Biotinylated antibody 2B7 showed purities >99% and a biotin incorporation rate of around 0.7, whereby the low value is favourable in preventing the formation of bead cross-links or aggregates. Labelling ratios ranging from 1:3.5–1:7.5 were tested for preparation of the Alexa Fluor® 647-labelled MW1 antibody. The highest labelling ratio delivered the highest incorporation rate and the highest fluorescence emission (**Supplemental Table 1**). A labelling ratio of 1:8 was used for production batches leading to an Alexa Fluor® 647 incorporation rate of 5.9 and a response/background signal ratio of >6 (**Supplemental Table 1**).

Suitability of the surrogate matrix was demonstrated by parallelism experiments in patient samples (**Supplemental Table 2**). Due to difficulty in obtaining large amounts of human CSF from healthy donors, artificial CSF (aCSF) stabilised with 1% Tween20 and a protease inhibitor was used as a surrogate matrix for the preparation of calibration standard and quality control (QC) samples. Suitability of the surrogate matrix was investigated by comparing the performance of the calibration curve in pooled human CSF and the surrogate matrix (**Supplemental Figure 1**). Using optimised capture and detection reagents together with the controlled assay matrix enhanced the assay signal-to-noise ratio at the lowest calibration (1.63 pg/mL HTT Q46) from 2–3 to 4–8 (assay pre-validation data, not shown).

After optimisation of pipetting and washing steps, an overall plate precision of the assay signals at a low QC (LQC) level of around 20% was reached and no systematic plate effects were observed. To mitigate low plate precision, all samples were analysed in triplicates to allow single-value outlier exclusion according to pre-defined criteria.

### Assessment of assay specificity and the adequacy of the reference standard

To assess the impact of HTT size or polyQ-repeat length on the assay signal, the concentration-response curves of 21 different recombinant HTTs were measured (**Figure 1A**). Assay signals across plates were normalised using repeated measures with the HTT Q45 protein, which led to highly similar concentration-responses (**Figure 1B**).

**Figure 1.**
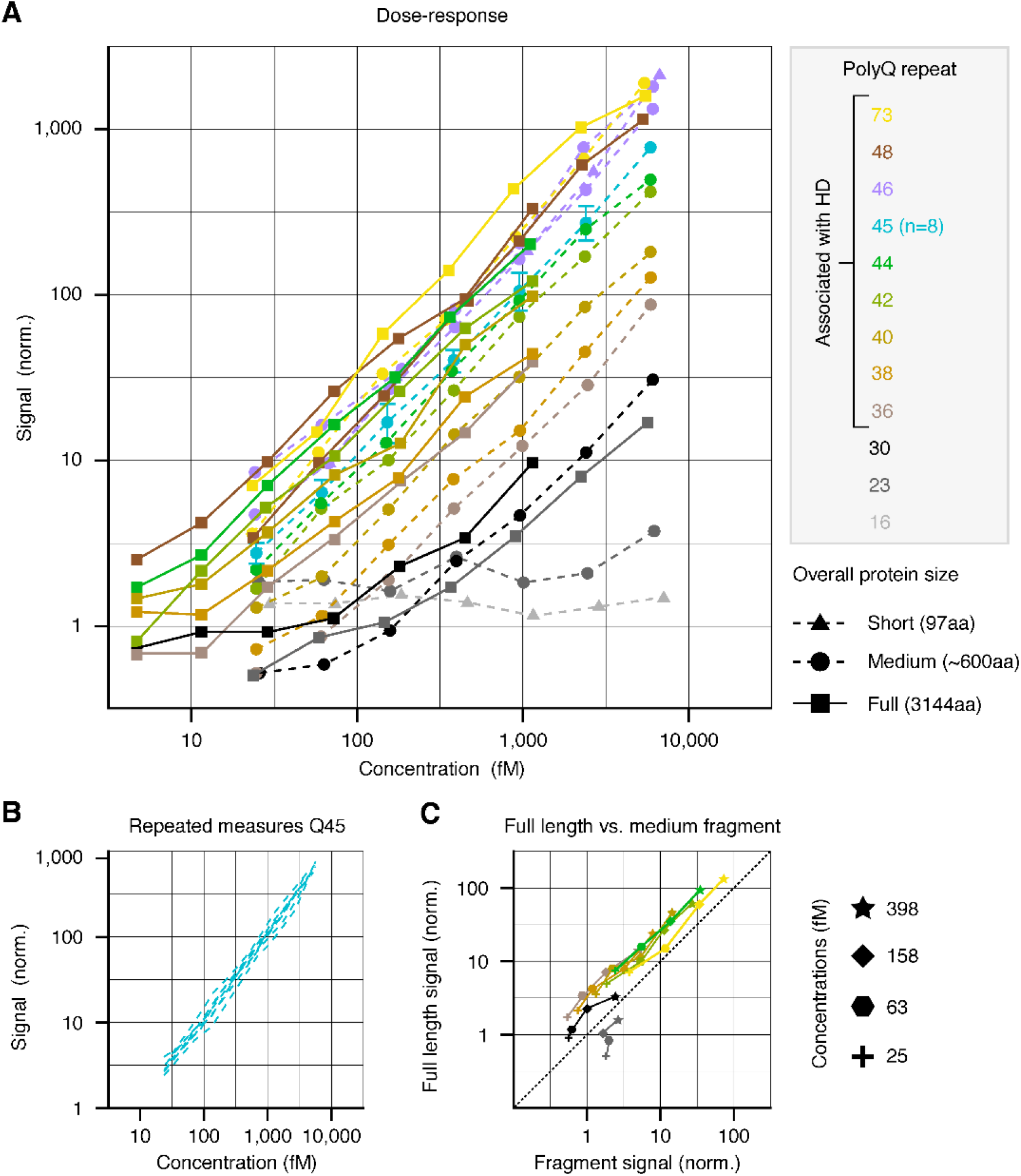
Dose-response of different recombinant HTT. (**A**) Normalised signals across recombinant proteins varying in the number of polyQ repeats (12 repeats tested) and overall protein size (three sizes tested). The number of polyQ repeats is indicated by colour while overall protein size is indicated by marker symbols. The protein with 45 polyQ repeats has been measured across all eight plates and the highest concentration was used to derive a normalisation factor applied to all dose-responses on the plate for cross-plate comparability. The error bars for the Q45 protein reflect the standard deviation across all eight runs. The Q46 medium size fragment was chosen as the reference protein during assay validation; both representative dose-responses depicted in the figure were measured separately. (**B**) All individual dose-responses of the Q45 protein measured across different plates. Note that normalisation induced the identical response at the highest concentration. (**C**) Normalised signals for fragment and full-length versions of proteins with the same polyQ repeat numbers and similar molarity. The same colour coding as for (**A**) was used to label the different polyQ repeat numbers. HD, Huntington’s disease; HTT, huntingtin protein; norm., normalisation; polyQ, polyglutamine.

A graded increase in the assay response was observed with increasing polyQ repeats and flat responses among the proteins with low wild-type levels of polyQ repeats (cf. Q16 and Q23 proteins, **Figure 1A**). As the number of polyQ repeats increased, an increase in assay response was observed, leading to robust dose-responses that were particularly notable for recombinant proteins with polyQ-repeat lengths associated with HD (>36). Among all proteins with ≥36 polyQ repeats, parallel, albeit shifted, concentration-responses were observed via log-log visualisation (**Figure 1A**). Interestingly, the steep signal gain observed from the wtHTT to the HTT with mid-40 polyQ repeats, i.e. vertical shift of the dose-responses, did not continue linearly towards very high polyQ repeats. The signal responses generated by recombinant proteins with 73 polyQ repeats partially overlapped with proteins in the mid-40 range, suggesting a saturation of signal response despite increasing polyQ repeats in that range.

To examine the role of concentration, overall protein size and polyQ-repeat length on the assay signal, a direct comparison was performed for four concentration levels and eight different polyQ-repeat lengths where signals under similar molarity were available for medium-size fragments and full-length proteins (**Figure 1C**). A three-way analysis of variance (ANOVA) confirmed that concentration, overall protein size and polyQ-repeat length all had significant main effects on the assay signal (all *P* < 10^−10^, ANOVA). Consistently higher signals were generated by the full-length proteins when compared with the medium-sized fragments (*P* < 10^−4^, signed rank test of the signal difference between full-length proteins and medium-sized fragments across all polyQ-repeat lengths and concentrations). In addition, longer polyQ repeats led to consistent increases in mHTT signal. Interestingly, the lowest polyQ-repeat length of 23 did not show a stronger signal for full-length protein when compared with fragment versions of the protein at the same molarity. However, a dose-response (i.e. increasing signal with increasing concentration) was present for the full-length but not the fragment protein. These observations suggest that cross-reactivity between MW1 and wtHTT can occur at extremely high, likely non-physiological protein concentrations and is facilitated by the presence of full-length HTT.

Following the observation of parallel dose-response curves for all recombinant proteins with ≥36 polyQ repeats (**Figure 1A**), the parallel nature of the dose-response curves was quantitatively assessed using linear mixed-effect models. Selection criteria for recombinant HTTs used in the modelling analyses required HTTs with polyQ-repeat lengths ≥36 and concentrations >26 fM (i.e. above the lower limit of quantification [LLOQ], within the working range of the assay).

Factors of influence in the analyses were the intercepts and slopes of the dose-response curves. Findings showed that the model, which assumed variations in intercept (null model, i.e. only vertical shifts in dose-response curves with identical slopes), delivered essentially equivalent predictions (r> 0.99) compared with a model that assumed variations in intercepts and slopes across proteins (test model, i.e. individual vertical shifts and slopes for each protein) (**Figures 2A, 2B and 2C**). However, a direct nested model comparison revealed that the test model fit the data significantly better than the null model (log-likelihood ratio test, *P* = 9 × 10^−4^; null model: Bayesian Information Criterion:

**Figure 2.**
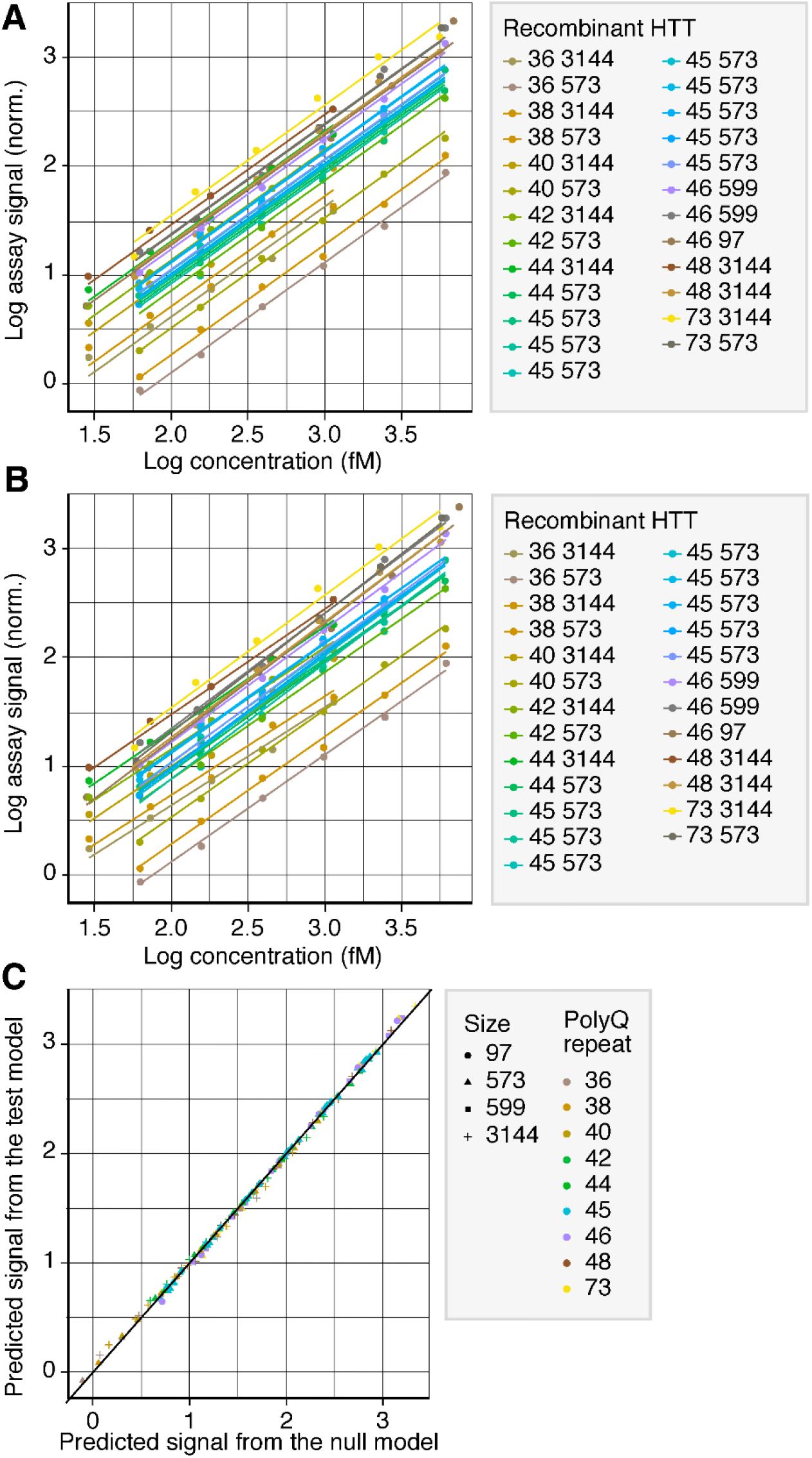
Model predictions of the null and test models across all dose-response curves. (**A**) The null model was defined by random intercepts and fixed slopes. (**B**) The test model was defined by random intercepts and random slopes. In (**A**) and (**B**) the legend indicates each recombinant HTTs’ polyQ-repeat length, overall length and plate number, respectively. Legends in (**A**) and (**B**) indicate each recombinant HTTs’ polyQ repeat length (first number) and protein size/number of amino acids (second number). (**C**) Comparison of predicted signals from the models in (**A**) and (**B**). Legend in (**C**) indicates the recombinant HTTs: icon shape denotes the protein size/number of amino acids; icon colour denotes the polyQ-repeat length. HTT, huntingtin protein; norm., normalisation; polyQ, polyglutamine.

–193.8, Akaike Information Criterion: –205.7; test model: Bayesian Information Criterion: – 197.8, Akaike Information Criterion: –215.7). We observed that the random slopes of the test model only represented a minor (typically <5%) variation around the overall slope for all proteins, suggesting the random slopes had a minimal impact on the overall model’s performance. Despite the minor impact of the random slopes on overall model predictions, the slopes correlated significantly and positively (Spearman rank correlation 0.73, *P* = 3.4 × 10^−5^) with polyQ repeats, suggesting slightly steeper dose-responses for proteins with higher polyQ repeat numbers. The correlation between polyQ-repeat length and slope was largely driven by proteins with <40 and >70 repeats, while proteins with mid-40 polyQ repeats varied more randomly from the overall slope both positively and negatively (e.g. see slopes for Q48 and Q42) (**Figure 3**). These findings showed that the null and test models performed equivalently, suggesting the dose-response curves across mutant versions of the recombinant HTTs were highly parallel.

**Figure 3.**
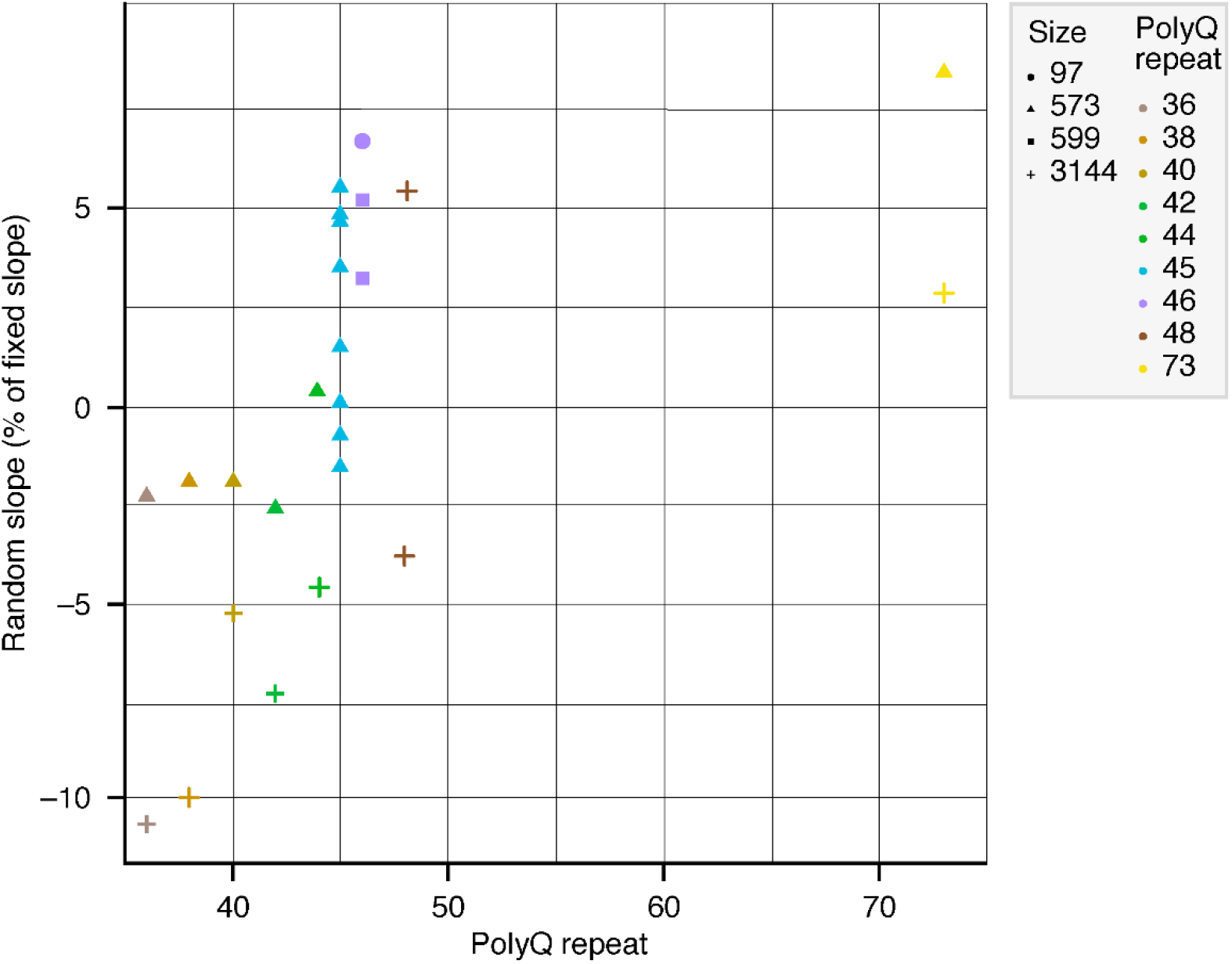
Relationship between polyQ-repeat length of recombinant HTTs and random slopes. Legends indicate the recombinant HTTs: icon shape denotes the protein size/number of amino acids; icon colour denotes the polyQ-repeat length. HTT, huntingtin protein; polyQ, polyglutamine.

To assess the effect of the reference-standard protein on the relative quantitation of CSF mHTT in a clinical trial setting, we performed simulations to evaluate how the slopes observed in the test model would affect concentration estimates in the assay if different proteins were used as reference standards.

Concentrations were back-calculated based on the different dose-response curves of the HTTs; two example assay signals were observed at a putative baseline (Log signal=1) and at a follow-up visit (Log signal=0.7), respectively, which simulated a decrease in CSF mHTT concentrations and resulted in a drop in the assay signal between the two visits (**Figures 4A–4D**).

**Figure 4.**
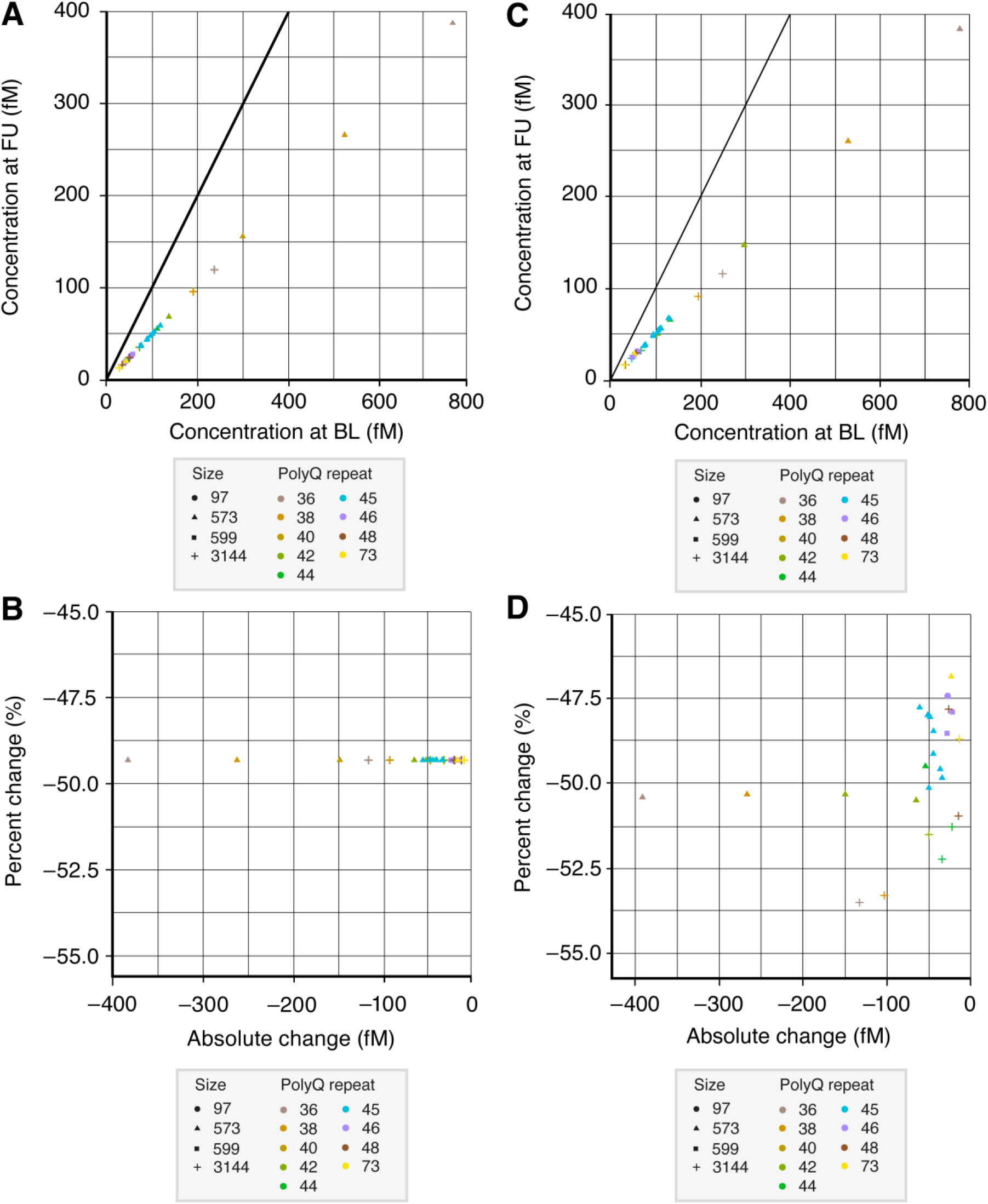
Simulation of longitudinal signals with an HTT-lowering effect. Two signal levels were used to simulate visits at baseline (Log signal=1) and at follow-up (Log signal=0.7), respectively. (**A**) The null model was defined by the random intercepts and fixed slopes that emerged from the null model fit to the dose-response curve data. (**B**) Percent change plotted against absolute change in the null model. (**C**) The test model was defined by the random intercepts and random slopes that emerged from the test model fit to the dose-response curve data. (**D**) Percent change plotted against absolute change in the test model. Legends outline the recombinant HTTs: icon shape denotes the protein size/number of amino acids; icon colour denotes the polyQ-repeat length. BL, baseline; FU, follow-up; HTT, huntingtin protein; polyQ, polyglutamine.

In both the null and test models, differences in the intercepts between the dose-response curves represented the largest source of variability between the recombinant HTTs, causing highly different estimated absolute concentrations. This means a protein with a higher intercept of the dose-response curve would yield lower concentration estimates than a protein with a lower intercept of the dose-response curve, even in the order of several hundred femto-molar, at the same observed assay signal. These observations emphasise the relative quantitative nature of the assay and the importance of the choice of reference standard (**Figures 4A and 4C**).

The null model yielded identical percent change signals across the different recombinant HTTs, due to its fixed slope (**Figure 4B**). This scaled baseline and follow-up visit concentration estimates to identical degrees, while keeping their ratio constant. In contrast, due to the variable slopes, the test model yielded small variations in the estimated percent change dependent on the reference protein. These variations were typically +/–3%, which were a minor fraction of the underlying mHTT-lowering signal simulations (**Figure 4D**). Wide variations in absolute change were observed for both the null and test models.

The simulated mHTT concentration change between baseline and follow-up visits was systematically varied in a physiologically meaningful range (∼60% decrease to 60% increase) between visits (**Figure 5**). The variability in the percentage change estimates between proteins was a function of the underlying mean percent change signal itself, whereby smaller percent changes in mHTT were associated with less variability across reference standards. This means the greater the mean fold change signal (decrease or increase), the higher the resulting variability of the fold change estimates derived from the different recombinant HTTs under the test model.

**Figure 5.**
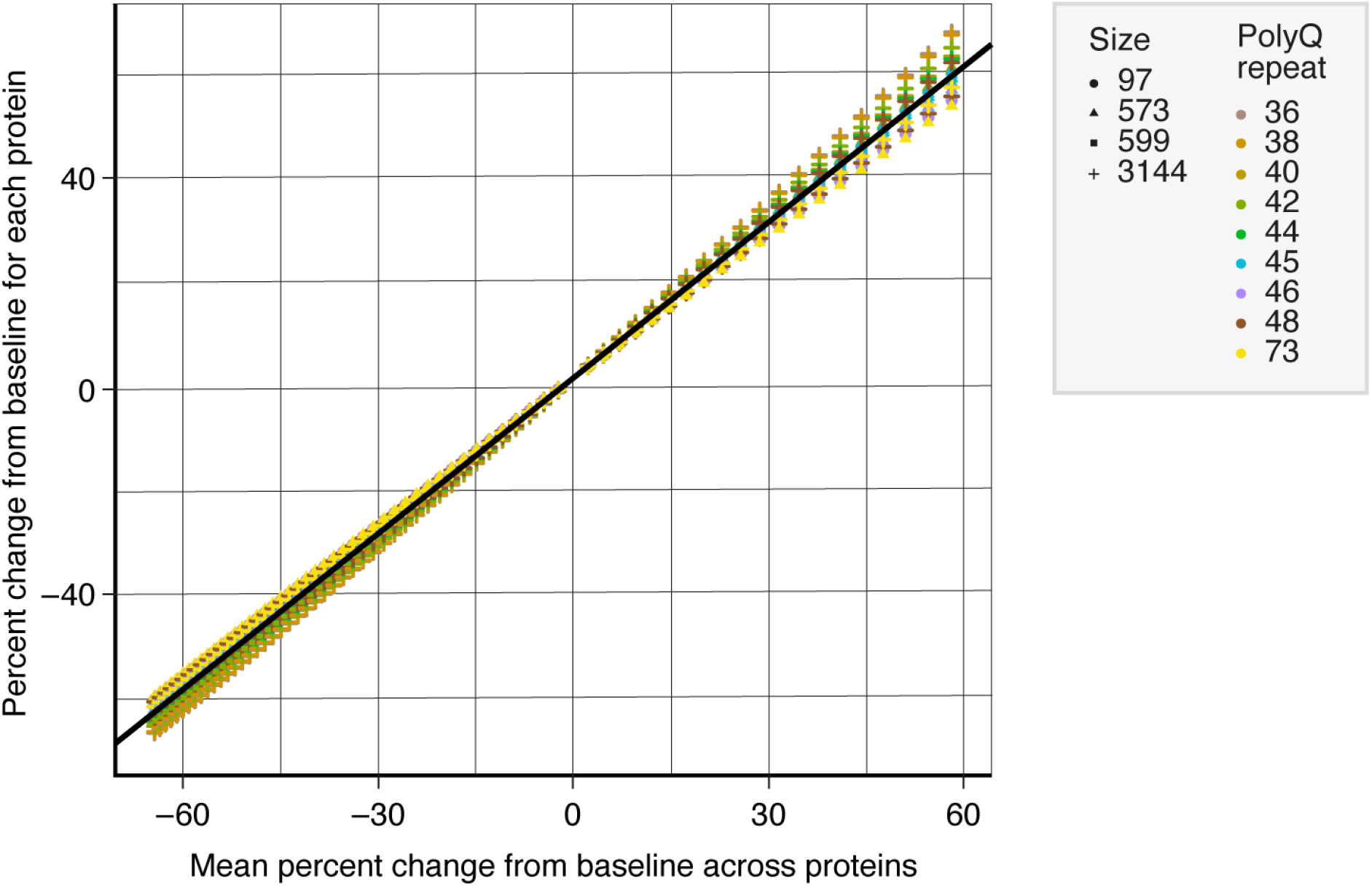
Variability of the estimated percent change signal in mHTT concentrations across recombinant proteins. Percent change from baseline for each recombinant HTT plotted against mean percent change from baseline across proteins. Legends indicate the recombinant HTTs: icon shape denotes the protein size (number of amino acids); icon colour denotes the polyQ-repeat length. HTT, huntingtin protein; mHTT, mutant HTT; polyQ, polyglutamine.

When the variability in the percentage change estimates (standard deviation) was normalised by the underlying signal change (mean) to derive the coefficient of variation, the fold change variability across recombinant HTTs constituted a minor fraction of the underlying change signal (in the order of 5% across all signal changes).

In sum, our observations suggest that there are minor variabilities between the concentration estimates based on different recombinant HTTs as the reference standard, although these variabilities are likely negligible compared with physiological signals of interest. As a consequence, any mid-40 polyQ-repeat HTTs may be suitable as a reference standard for the assay and deliver equivalent results for clinical trials, in the context of relative quantitation (i.e. while considering patient data as relative to the patients’ baseline value). The Q46 medium size fragment was used for assay validation, in keeping with previous research-grade versions of the assay. We expect the choice of a different reference standard with similar properties (e.g. a Q42 fragment or Q48 full length) to alter absolute concentration estimates but to deliver highly consistent patterns of relative change. The same pattern should hold true across patients with different polyQ-repeat lengths whose CSF mHTT levels are quantified against a particular reference standard that may not represent the polyQ length of that patient. Different reference standards will therefore deliver the same pattern of mHTT change across a range of patients with differing polyQ-repeat lengths. A caveat may emerge for patients with very low numbers of polyQ repeats where it may be more empirically difficult to detect HTT in the CSF of patients due to the lower assay signal delivered by low polyQ-repeat HTTs.

### Assay validation

#### Performance of HTT Q46 calibrators during method validation

Validation in two independent laboratories confirmed the assay had high sensitivity. The calibration range was 1.63 pg/mL (LLOQ – Roche) and 0.655 pg/mL (anchor point – ICON [validated LLOQ: 1.64 pg/mL]), to 400 pg/mL (upper limit of quantification [ULOQ]) HTT Q46 in surrogate matrix, determined via parallelism data. HTT Q46 calibrators prepared with reference standard spiked in surrogate matrix performed well during method validations (**Table 1**). Accuracy of all individual calibration samples were within the acceptance criteria of 70%–130% accuracy. The calibrators enabled full recovery of frozen QC samples.

**Table 1.** Precision and accuracy of calibration standards. Back-calculated mHTT concentrations (pg/mL) for calibration standards in surrogate matrix.

| Nominal HTT Q46 concentrations in surrogate matrix (pg/mL) |  |  |  |  |  |  |  |  |  |  |  |  |  |  |
| --- | --- | --- | --- | --- | --- | --- | --- | --- | --- | --- | --- | --- | --- | --- |
|  | Roche | ICON | Roche | ICON | Roche | ICON | Roche | ICON | Roche | ICON | Roche | ICON | Roche | ICON |
|  | 1.63 | 1.64 | 4.08 | 4.10 | 10.2 | 10.2 | 25.6 | 25.6 | 64.0 | 64.0 | 160 | 160 | 400 | 400 |
| Mean | 1.62 | 1.67 | 4.16 | 4.07 | 10.3 | 10.3 | 24.8 | 25.2 | 65.5 | 67.1 | 161 | 156 | 402 | 409 |
| Precision (%CV) | 1.5 | 6.5 | 4.3 | 4.7 | 3.0 | 4.3 | 4.6 | 4.5 | 4.8 | 6.9 | 6.0 | 6.9 | 4.4 | 6.8 |
| Relative error / bias (%) | −0.6 | 1.7 | 2.0 | −0.7 | 1.0 | 1.0 | −3.1 | −1.4 | 2.3 | 4.8 | 0.6 | −2.6 | 0.5 | 2.3 |
| n | 18 | 24 | 18 | 25 | 18 | 25 | 18 | 25 | 18 | 25 | 18 | 25 | 18 | 25 |

CV, coefficient of variation; HTT, huntingtin protein.

#### Intra- and inter-assay accuracy and precision in spiked surrogate matrix

Overall, inter- and intra-assay accuracy and precision of the reference standard (HTT Q46) in surrogate matrix (aCSF) matched the predefined acceptance criteria across both laboratories (**Table 2**). In both validations, the determined mean concentration at each level (including the LLOQ and ULOQ) was within 70%–130% accuracy; precision of the mean concentration determined at each level was ≤30% coefficient of variation (CV).

**Table 2.** Intra- and inter-assay accuracy and precision in spiked surrogate matrix.

|  | LLOQ (pg/mL) |  | LQC (pg/mL) |  | MQC (pg/mL) |  | HQC (pg/mL) |  | ULOQ (pg/mL) |  |
| --- | --- | --- | --- | --- | --- | --- | --- | --- | --- | --- |
|  | Roche | ICON | Roche | ICON | Roche | ICON | Roche | ICON | Roche | ICON |
|  | 1.63 | 1.64 | 4.50 | 4.50 | 40.0 | 40.0 | 300 | 300 | 400 | 400 |
| Intra-assay |  |  |  |  |  |  |  |  |  |  |
| Intra-assay accuracy (%) | 84.7 | 100.3–119.7 | 71.8 | 100.2–115.5 | 79.0 | 97.4–120.2 | 73.3 | 80.5–115.8 | 79.3 | 80.8–120.0 |
| Intra-assay precision (%CV) | 14.1 | 14.1 | 12.1 | 5.6 | 6.2 | 4.0 | 4.1 | 6.2 | 6.7 | 12.9 |
| Total error (%) | 29.4 | NA | 40.3 | NA | 27.2 | NA | 30.8 | NA | 27.4 | NA |
| Inter-assay |  |  |  |  |  |  |  |  |  |  |
| Inter-assay accuracy (%) | 93.3 | 109.5 | 77.6 | 106.2 | 84.8 | 106.9 | 84.0 | 99.3 | 83.8 | 93.3 |
| Inter-assay precision (%CV) | 11.5 | 0.00 | 9.5 | 4.7 | 5.4 | 7.5 | 9.4 | 11.1 | 7.8 | 13.3 |
| Total error (%) | 18.2 | NA | 31.9 | NA | 20.6 | NA | 25.4 | NA | 24.0 | NA |
| Intra-assay and Inter-assay |  |  |  |  |  |  |  |  |  |  |
| Overall bias (%) | NA | 9.5 | NA | 6.2 | NA | 6.9 | NA | -0.7 | NA | -6.7 |
| Total precision (%CV) | NA | 14.1 | NA | 7.3 | NA | 8.5 | NA | 12.7 | NA | 18.5 |
| Total error** (%) | NA | 23.6 | NA | 13.5 | NA | 15.4 | NA | 13.4 | NA | 25.2 |

\* Sum of absolute value of bias and precision.
\*\* Sum of absolute value of overall bias and total precision.
CV, coefficient of variation; HQC, high quality control; LLOQ, lower limit of quantification;
LQC, lower quality control; MQC, mid quality control; ULOQ, upper limit of quantification.

#### Inter-assay precision in the CSF of patients with HD

Precision of CSF samples from patients with HD were reliable across multiple independent runs within each laboratory as well as across the independent laboratories (**Table 3**). In both validations, the precision of the mean concentration determined for each patient sample met the acceptance criteria of ≤30% CV precision.

**Table 3.**
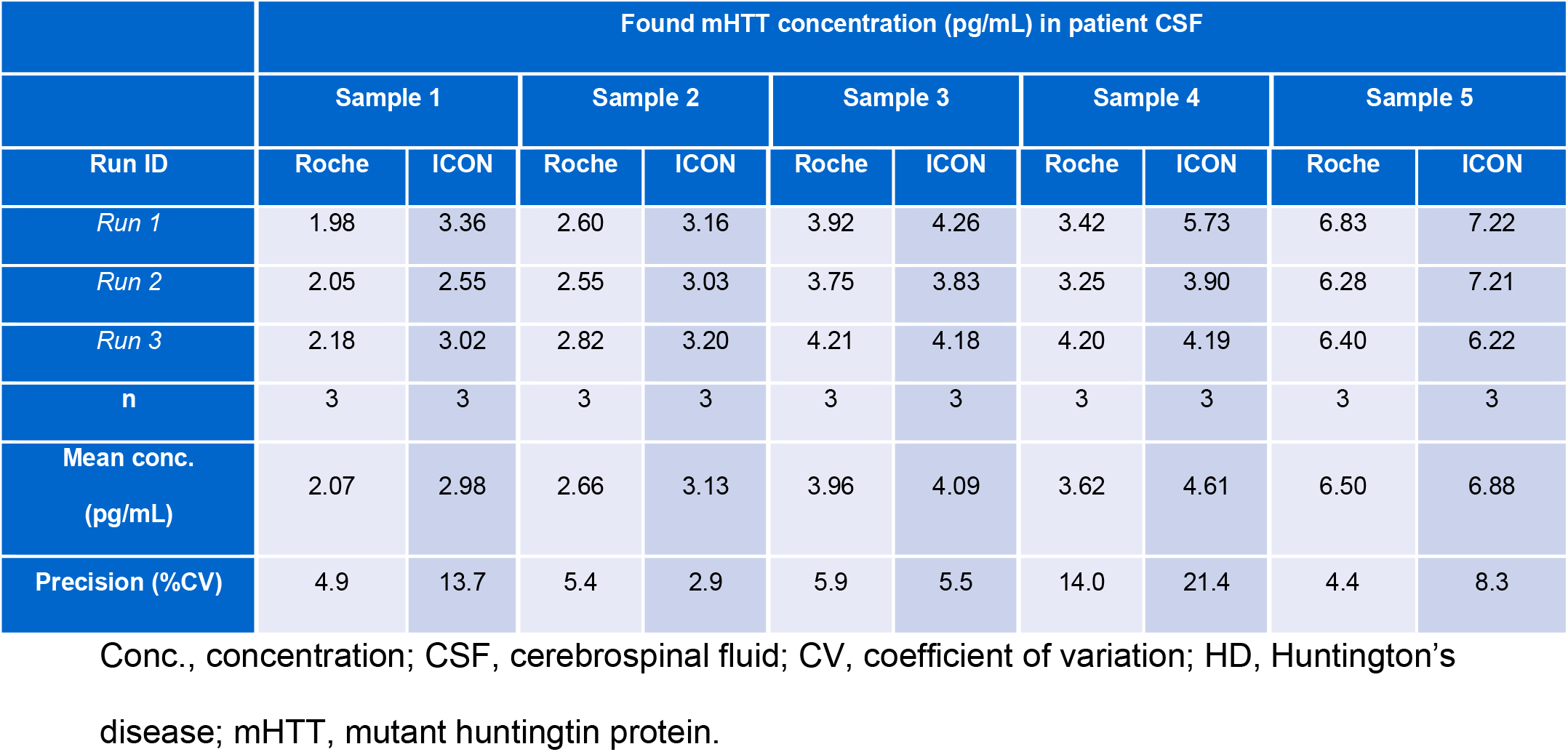
Inter-assay precision in the CSF of patients with HD.

#### Parallelism

To confirm comparable behaviour of different HTTs in patient CSF as the real matrix of interest, the assay signal of real patient CSF with largely differing polyQ-repeat lengths were measured (tested polyQ-repeat lengths: 41, 42, 44, 48, 50, 51) in serial dilution with surrogate matrix using the Q46 reference standard to back-calculate concentration estimates. Dilutional parallelism was demonstrated in all samples, including patients with low polyQ-repeat lengths (polyQ repeats 41, 42 [**Supplemental Table 3**]; polyQ repeats 44–51 [**Table 4**]), further indicating a comparable assay signal behaviour across the spectrum of HD-related HTTs. The precision of the mean concentration across all dilutions within the dynamic assay range was ≤16.7% across both validations, fulfilling the acceptance criteria of ≤30% CV (**Table 4**). The parallelism data (**Table 4)** were used to determine the parallelism LLOQ (LLOQ_P_). Comparable values of 1.57 pg/mL (Roche) and 1.69 pg/mL (ICON) were obtained.

**Table 4.** Parallelism data for mHTT in the CSF of patients with HD. Serial dilution of samples from patients with mid–long polyQ-repeat lengths in surrogate matrix.

|  |  | Roche |  |  | ICON |  |  |
| --- | --- | --- | --- | --- | --- | --- | --- |
| Sample | Dilution factor | Back-calculated conc.<br>(pg/mL) | Undiluted<br>conc.<br>(pg/mL) | Recovery<br>(%) | Back-calculated<br>conc.<br>(pg/mL) | Undiluted<br>conc.<br>(pg/mL) | Bias (%) |
| 1 | 1 | 4.26 | 4.68 | 100 | 5.50 | 5.50 | 0 |
|  | 2 | 2.40 | 5.29 | 113 | 2.63 | 5.26 | –4.3 |
|  | 3 | 1.57* | 5.19 | 110.9 | 1.64 | 4.91 | –10.7 |
|  | 4 | 1.56 | 6.87 | NA | 1.69* | 6.78 | 23.2 |
|  | 6 | 0.601 | 3.96 | 84.6 | NV | NA | NA |
|  | 8 | 0.403 | 3.55 | 75.9 | 0.167 | 1.33 | **–75.7** |
| Mean (pg/mL) |  | 5.05 |  |  | 5.61 |  |  |
| CV (%) |  | 6.5 |  |  | 14.5 |  |  |
| n (results within curve) |  | 3 |  |  | 4 |  |  |
| 2 | 1 | 5.07 | 5.57 | 100 | 6.96 | 6.96 | 0 |
|  | 2 | 2.53 | 5.57 | 100 | 3.45* | 6.89 | –1.0 |
|  | 3 | 1.79 | 5.90 | 105.9 | 1.60 | 4.79 | **–31.1** |
|  | 4 | 1.11 | 4.86 | 87.3 | 1.72 | 6.89 | –0.9 |
|  | 6 | 0.762 | 5.03 | 90.3 | 1.02 | 6.11 | –12.1 |
|  | 8 | 0.521* | 4.58 | 82.2 | 0.625 | 5.00 | –28.1 |
| Mean (pg/mL) |  | 5.68 |  |  | 6.38 |  |  |
| CV (%) |  | 3.4 |  |  | 16.6 |  |  |
| n (results within curve) |  | 3 |  |  | 4 |  |  |
| 3 | 1 | 5.20 | 5.72 | 100 | 4.86 | 4.86 | 0 |
|  | 2 | 2.45 | 5.40 | 94.4 | 1.97 | 3.95 | –18.9 |
|  | 3 | 1.55*, a | 5.12 | 89.5 | 1.53 | 4.59 | –5.7 |
|  | 4 | 0.856 | 3.77 | **65.9** | 1.10 | 4.39 | –9.7 |
| Sample | Dilution factor | Back-calculated conc.<br>(pg/mL) | Undiluted<br>conc.<br>(pg/mL) | Recovery<br>(%) | Back-calculated<br>conc.<br>(pg/mL) | Undiluted<br>conc.<br>(pg/mL) | Bias (%) |
|  | 6 | 0.749 | 4.94 | 86.4 | 0.646* | 3.88 | -20.3 |
|  | 8 | 0.454 | 3.99 | **69.8** | 0.803 | 6.43 | **32.1** |
| Mean (pg/mL) |  | 5.41 |  |  | 4.47 |  |  |
| CV (%) |  | 5.5 |  |  | 10.5 |  |  |
| n (results within curve) |  | 3 |  |  | 3 |  |  |
| 4 | 1 | 5.73 | 6.30 | 100 | 3.50 | 3.50 | 0 |
|  | 2 | 2.83 | 6.23 | 98.9 | 1.60 | 3.20 | -8.3 |
|  | 3 | 2.20 | 7.27 | 115.4 | 1.01 | 3.04 | -13.1 |
|  | 4 | 1.84 | 8.11 | 128.7 | 0.800* | 3.20 | -8.5 |
|  | 6 | 0.905 | 5.97 | 94.8 | 1.168 | 7.01 | **100.5** |
|  | 8 | 0.633* | 5.57 | 88.4 | 0.410 | 3.28 | -6.2 |
| Mean (pg/mL) |  | 6.98 |  |  | 3.35 |  |  |
| CV (%) |  | 12.8 |  |  | 6.1 |  |  |
| n (results within curve) |  | 4 |  |  | 2 |  |  |
| 5 | 1 | 8.58 | 9.44 | 100 | NV | NA | NA |
|  | 2 | 4.21 | 9.26 | 98.1 | 2.69 | 5.38 | 0 # |
|  | 3 | 2.57 | 8.48 | 89.8 | 1.77 | 5.32 | -1.2 # |
|  | 4 | 2.14 | 9.42 | 99.8 | 1.47 | 5.89 | 9.4 # |
|  | 6 | 1.20 | 7.91 | 83.8 | 0.699* | 4.20 | -22.1 # |
|  | 8 | 1.05* | 9.26 | 98.1 | NV | NA | NA |
| Mean (pg/mL) |  | 9.15 |  |  | 5.53 |  |  |
| CV (%) |  | 5.0 |  |  | 5.6 |  |  |
| n (results within curve) |  | 4 |  |  | 3 |  |  |
| Sample | Dilution factor | Back-calculated conc.<br>(pg/mL) | Undiluted<br>conc.<br>(pg/mL) | Recovery<br>(%) | Back-calculated<br>conc.<br>(pg/mL) | Undiluted<br>conc.<br>(pg/mL) | Bias (%) |
| 6 | 1 | 5.95 | 6.54 | 100 | 4.25 | 4.25 | 0 |
|  | 2 | 2.73 | 6.01 | 91.9 | 1.91 | 3.81 | -10.2 |
|  | 3 | NV | NA | NA | 1.46* | 4.39 | 3.3 |
|  | 4 | 1.41a | 6.20 | 94.8 | 1.39 | 5.57 | **31.3** |
|  | 6 | 0.915 | 6.04 | 92.4 | 0.681 | 4.08 | -3.8 |
|  | 8 | 0.574* | 5.05 | 77.2 | 0.408 | 3.26 | -23.1 |
| Mean (pg/mL) |  | 6.25 |  |  | 4.50 |  |  |
| CV (%) |  | 4.3 |  |  | 16.7 |  |  |
| n (results within curve) |  | 3 |  |  | 4 |  |  |

\*Lowest concentration for each sample providing a parallel response (i.e. recovery within 70%–130%).
#, Because a triplicate CV >20.0% was observed in the undiluted sample, the first 2-fold dilution result was used as a reference to calculate the bias.

#### Microplate homogeneity

Appropriate microplate homogeneity was demonstrated at the LLOQ level since the accuracies for all mimicry LLOQ samples were between 72.5% and 122.1% (**Supplemental Table 4**).

#### Interferences of wtHTT, drug and blood

No relevant interferences of wtHTT (**Supplemental Table 5**) or drug (**Supplemental Table 6**) were shown in the assay. Interference of blood differed between the two independent validations. One laboratory observed interference at 1.0% whole blood in samples analysed and spiked at 1.64 pg/mL. In contrast, data from the other independent laboratory fulfilled the acceptance criteria for the absence of interference of blood (**Supplemental Table 7**).

#### Prozone effect

No high-dose hook effect was observed up to the highest-tested concentration of 12.5 μg/mL of tominersen. The 1/100,000 dilution generated a result within the working range with an accuracy of 91.2%.

#### Stability of reference standard in surrogate matrix

Bench-top stability of the reference standard in surrogate matrix was demonstrated for at least 4 hours (**Supplemental Table 8**). Stability of the reference standard in surrogate matrix was demonstrated for up to 12 months of storage at –60 °C to –85 °C (**Supplemental Tables 9 and 10**).

## Discussion

With the ongoing development of HTT-lowering therapies for HD, changes in the levels of CSF mHTT may be a critical biomarker that may capture a biological signal with direct causal relevance in the trajectory of HD pathology. To support regulatory decision-making processes in drug development, it is important to ensure biomarker assays are both robust and reliable. Furthermore, these assays should comply with international regulatory guidelines while maintaining transferability and generating replicable data.

The current study performed validations in two independent laboratories, aimed at generating a bead-based sandwich ligand binding assay that fulfils regulatory requirements as depicted in regulatory guidelines. Good Laboratory Practice is an example of Good Practice guidelines – a collection of international regulations established to ensure the quality standard of products in multiple fields, such as pharmacological drug development (14). Additional analyses were also performed in this study to further characterise the assay methodology. Translation of the mHTT ligand binding assay from a research-grade environment to regulated validation in clinical-grade laboratories enables the assay to serve as a valuable resource that will facilitate the clinical development of HTT-lowering therapies.

Comparison of the assay signal across a wide range of recombinant HTTs showed a steep increase in assay signal, with the transition of HTT from non-disease causing (<36 polyQ repeats) to definitively disease causing (≥40 polyQ repeats). All tested recombinant mHTTs delivered robust dose-responses that were highly parallel, supported by quantitative linear mixed-effect modelling data on multiple recombinant proteins with polyQ-repeat lengths ≥36. This was irrespective of polyQ length, overall protein size or expression system (**Table 5**).

**Table 5.**
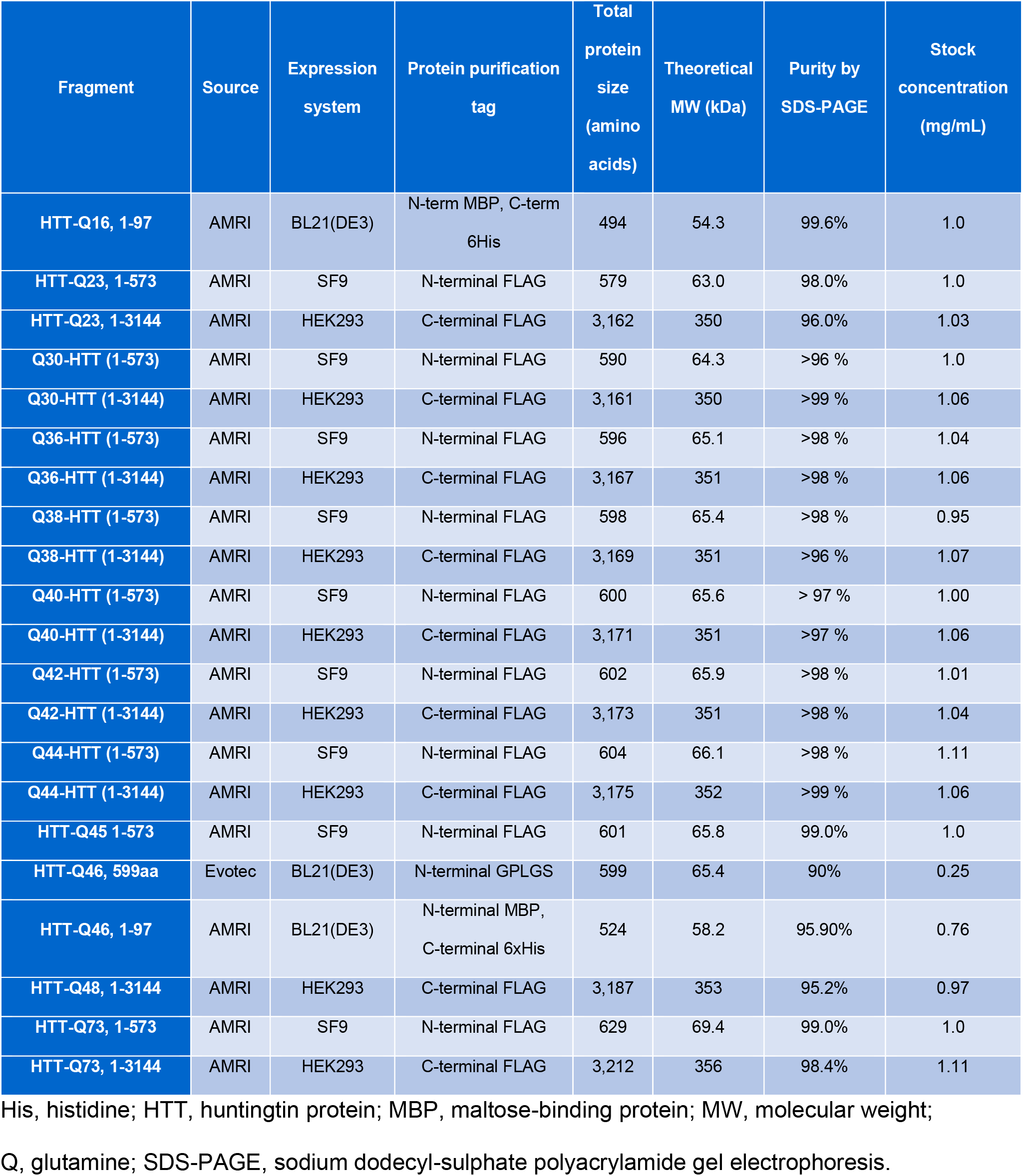
Overview of all HTT fragments.

These observations suggest that the assay reported here is broadly relevant across the HD continuum, with comparable signal properties across a wide range of polyQ repeats. Overall, the pre-set criteria for validation parameters were met, fulfilling the requirements for precision and accuracy in spiked surrogate matrix; precision in CSF from individuals with HD; parallelism; specificity; prozone effect and microtiter plate homogeneity. Parallelism data also demonstrated the absence of a matrix effect, which refers to any impact of assay components, aside from the analyte, on the analytical properties of the assay (15). As parallelism experiments generate curves that represent the binding affinity of the analyte as well as interference from the matrix, the presence or absence of a matrix effect can be inferred from the success or failure of a parallelism experiment (16).

These validation findings support the reliability of this ultra-sensitive bioanalytical method for quantifying mHTT in human CSF and show that it can be replicated and transferred. Notably, given that this is now a state-of-the art clinical-grade assay, previous findings on the levels of CSF mHTT in individuals with HD that were generated via the research-grade version of the assay may differ from future data.

A limitation of the assay is its relative quantitative nature, requiring the choice of a particular reference standard against which heterogeneous patient samples are being compared. As a result, absolute concentrations that are estimated with this method may exhibit inter-patient variability of a technical rather than biological nature. In future experiments it will be important to disentangle technical and biological variance of the assay signal in greater detail cross-sectionally, to evaluate the assay’s full scope as biomarker tool. Despite the limitations of requiring a particular reference standard, absolute concentrations from this assay may potentially carry valuable information about individual’s disease burden and trajectory when modelled appropriately in large data sets.

A further limitation is that sampling large amounts of CSF from healthy volunteers presents practical and ethical difficulties. As such, aCSF was used as a surrogate for human CSF during the preparation of calibrator and QC samples. The controlled environment afforded by the surrogate matrix is an important element for reliable assay performance, as justified by parallelism data.

This bead-based sandwich ligand binding assay developed for the quantification of changes in mHTT levels in human CSF has been successfully characterised and independently validated in two laboratories. Our findings show that this assay may be a reliable tool for generating biomarker data in registrational clinical trials for HD, with relevance across the HD continuum. Collaboration within the HD community will enable further refinement and application of this assay, supporting the development of HTT-lowering therapies for HD.

## Methods

### Materials

HTT fragments of different polyQ-repeat length and protein fragment size were purchased from AMRI and Evotec. Evotec HTT Q46, 599aa, was selected as the reference standard, and the surrogate matrix was purchased from Bio-Techne. Tween20 and protease inhibitor (cOmplete Protease Inhibitor™ Cocktail) were purchased from Sigma-Aldrich and Roche, respectively. The capture antibody was purchased from Evotec, and the detection antibody was expressed at Roche. SMC™ Capture Antibody Labeling kit (Merck Millipore) was used. Alexa Fluor® 647 carboxylic acid succinimidyl ester was purchased from Thermo Fisher Scientific. The centrifugal plate washer Blue® Washer (BlueCatBio, Germany) was used for plate wash steps. SMCxPRO™ specific buffers and glass-bottom 384-well plates were purchased from Merck Millipore. Ninety-six well polypropylene V-bottom plates were purchased from Brooks Life Sciences.

All human CSF samples were derived from individuals with early-manifest HD, obtained from the open-label extension (OLE) of the Phase I/IIa study of tominersen (NCT03342053).

### Assay setup

All results were generated by Good Clinical Practice-trained personnel in a regulated bioanalytical environment with Good Laboratory Practice-certified laboratories. A bead-based sequential ligand binding assay with SMC detection was used on the SMCxPRO™ (Merck) platform.

The ultra-sensitive assay employs the antibody pair 2B7/MW1 for capture and detection (**Figure 6**) and aCSF as a surrogate matrix. Capture antibody 2B7 binds to the N17 region of HTT (i.e. binds to both mHTT and wtHTT) and conjugates to streptavidin-coated magnetic particles via biotin coupling. Detection antibody MW1 is specific to the polyQ stretch present in mHTT and was labelled with Alexa Fluor® 647.

**Figure 6.**
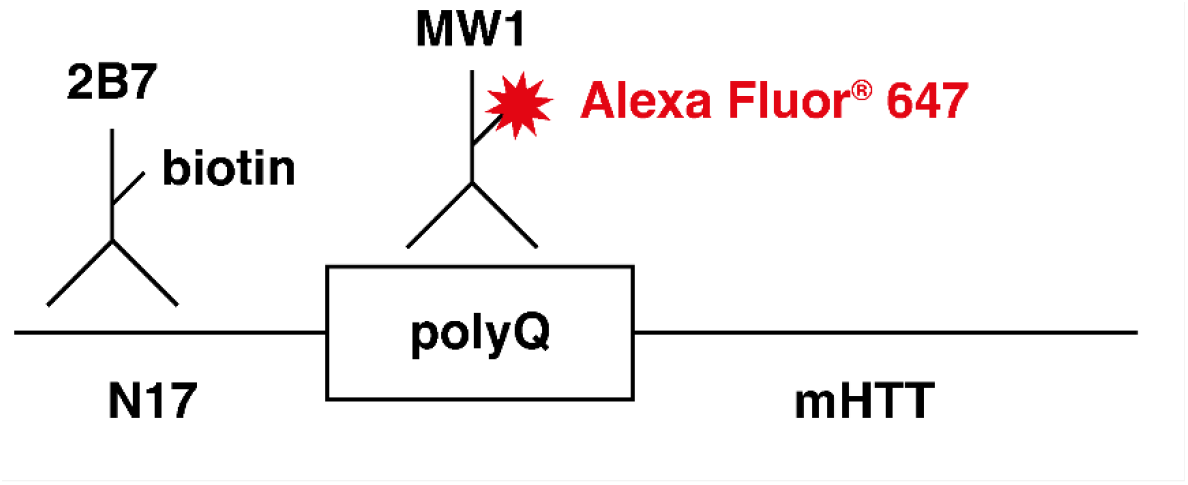
mHTT bead-based ligand binding assay: capture and detection of antibody-binding regions. mHTT, mutant huntingtin protein; polyQ, polyglutamine.

A 599 amino acid-long recombinant HTT fragment containing a Q46 amino acid-long polyQ chain was used as the reference standard (HTT Q46, molecular weight 65390 g/mol). Calibration standard and QC samples were prepared in surrogate matrix containing 1% Tween20 and a protease inhibitor cocktail (surrogate matrix) for an assay range of 1.63 pg/mL–400 pg/mL. The minimum required dilution (MRD) of the assay was set to 2.

### Optimisation of assay reagent preparation

Labelling of capture and detection antibodies were prepared at Roche Diagnostics (Penzberg, Germany), and labelled antibodies were analytically characterised at Roche Pharma Research (Penzberg, Germany). Antibody 2B7 was biotinylated using reagents and instructions from the SMC™ capture reagent labelling kit. The conjugate was purified via Superdex® 200 size exclusion chromatography. Coupling of biotinylated 2B7 to magnetic beads was performed according to kit instructions. MW1 was coupled with Alexa Fluor® 647-NHS ester with challenge ratios ranging from 1:3.5–1:8. Purification was performed via cut-off filtration (40K MWCO Zeba™ Spin columns). Spectrophotometry was used to determine the antibody concentration and Alexa Fluor® 647 labelling rate. The purity was determined by size exclusion-high-performance liquid chromatography, and the fluorescence emission level was determined by fluorescence spectrophotometry. Labelled antibodies and coupled beads were stored at 2–8 °C.

### Assay protocol

All calibration standards, QC samples and unknown samples were measured in triplicate wells. QC samples were prepared by spiking the reference standard in surrogate matrix, followed by shock freezing on dry ice and storage at or below –65 °C. Calibration samples were prepared on the day of the assay by spiking the reference standard in surrogate matrix. All buffers and reagents were equilibrated to room temperature before use and all assay steps were performed at room temperature.

A blocking buffer (50 μL/well) was dispensed into a 96-well V-bottom polypropylene plate, followed by either 150 μL/well calibration standards or a 15 μL/well sample dilution buffer containing 10% Tween20 and 1x protease inhibitor, and a QC or study sample (135 μL/well). Coupled beads (100 μL/well) diluted 1:500 in an Assay Discovery Buffer were added to the plate using a 12-channel manual pipette. The plate was sealed and incubated for 1.5 hours while shaken at 400 rpm. After incubation, the plate was placed for 2 minutes on the magnet of the Blue® Washer before centrifugation at 800 rpm. A sterile filtered detection antibody (20 μL/well) diluted 1:1000 in assay buffer was immediately added to the plate and incubated for 1 hour in a shaker at 700 rpm. After incubation, the plate was placed for 2 minutes on the magnet of the Blue® Washer before performing four wash cycles at 800 rpm with a 200 μL System Buffer added at each step. The plate was incubated with the last wash buffer for 2 minutes in a shaker at 700 rpm and the solution was transferred to a second microplate using a 12-channel manual pipette. The plate was incubated for 2 minutes on the magnet of the Blue® Washer and the plate was centrifuged at 800 rpm to remove the buffer. An elution buffer (12 μL/well) (Buffer B) was added to each well and the plate was placed in a shaker at 700 rpm for 6 minutes (performed using the Hamilton MicroLab Starline at ICON). A neutralisation buffer (10 μL/well) (Buffer D) was added to the glass-bottom 384-well reading plate using a manual 12-channel pipette (Roche) and the Hamilton MicroLab Starline (ICON). After placing the 96-well assay plate on a magnet for 2 minutes minimum, the supernatant (10 μL/well) was transferred from the 96-well assay plate to the 384-well reading plate using a manual 12-channel pipette (Roche) and the Hamilton MicroLab Starline (ICON). The 384-well reading plate was sealed with adhesive aluminium foil, placed in a shaker for 2 minutes at 700 rpm and spun for 5 minutes at 500 g. The plate was then placed on the bench for 30 minutes before readout on the SMCxPRO™ platform.

Data analysis was performed with Watson LIMS software (Roche) and Softmax Pro GxP 6.4 (ICON). For each triplicate well, the mean signal, standard deviation and the precision (%CV) were calculated. The calibration standards were fitted with a 4-parameter logistic with a weighting factor of 1/mean signal^2^. Concentrations of mHTT in samples were back-calculated using the fitted function and a minimum dilution factor of 1.1. Results of study samples showing signals below the LLOQ were reported as BLQ, provided they were measured at MRD.

### Assay validation

Validation parameters and acceptance criteria were adapted to the context of use and to the assay performance observed during pre-validation experiments. All samples were analysed in triplicates and the mean assay signal reported if the triplicate precision was ≤20%. Due to the difficulty in obtaining sufficient human CSF from healthy donors, assay validations in both independent laboratories were performed using the surrogate matrix.

A calibration standard curve was developed, consisting of seven non-zero calibration standards covering the dynamic assay range (1.63/1.64 pg/mL–400 pg/mL). The LLOQ and ULOQ were defined as the lowest and highest calibration standard concentrations within the dynamic range, respectively. Acceptance criteria required a minimum of six non-zero calibrator levels to have an accuracy of 70%–130%.

Inter-assay accuracy and precision in spiked surrogate matrix were assessed using a calibration standard curve and three sets of QC samples at the following concentrations: LQC (4.50 pg/mL), mid QC (40.0 pg/mL), high QC (HQC, 300 pg/mL) plus LLOQ/ULOQ samples. Intra-assay accuracy and precision were assessed using a calibration standard curve and three (ICON) or four (Roche) sets of QC samples at the five concentrations mentioned above for the inter-assay assessment. Measurements for inter- and intra-assay accuracy and precision were recorded in six independently prepared runs. Acceptance criteria for inter- and intra-assay accuracy and precision in spiked surrogate matrix required the determined mean concentration at each level including LLOQ and ULOQ to be within 70%–130% accuracy; precision of the mean concentration determined at each level needed to be ≤30% CV from the LLOQ to the ULOQ; and the total error needed to be ≤40%. Inter-assay precision was also assessed for CSF samples from patients with HD, where five patient samples measured in three independently prepared runs were performed on three different days. Acceptance criteria required the precision of the mean concentration determined at each level to be ≤30% CV from the LLOQ to the ULOQ.

Parallelism was assessed by the analysis of six samples from patients with HD. Study samples were serially diluted with surrogate matrix down to the LLOQ and below.

Recoveries were calculated based on concentration of the sample diluted at the MRD. Acceptance criteria for parallelism experiments required precision of the mean concentration across all dilutions within the dynamic assay range to be ≤30% CV for at least five out of the six tested samples. LLOQ_P_ was determined via parallelism data using the common concentration method (17) on data from six individuals with HD. Parallelism data were also used to validate the MRD.

Microplate homogeneity was assessed by adding a full set of calibration standards and QC samples, prepared in surrogate matrix, to an analytical run. A single volume of an LLOQ sample in surrogate matrix sufficient to fill all remaining free positions for validation samples of an analytical run (excluding calibration standards and analytical run acceptance QC samples) was prepared and added. The LLOQ sample was quantified using triplicate mean evaluation of study samples. Acceptance criteria for microplate homogeneity required ≥80% of the LLOQ samples to show accuracies within 70% and 130%.

Potential interference of wtHTT was assessed using a full-length HTT containing a Q23 polyQ chain (HTT-Q23 1-3144 aa) spiked at 0, 20.0, and 200.0 pg/mL (corresponding to 0,

47.5 and 475.0 fM) in blank surrogate matrix and in surrogate matrix spiked at LLOQ and HQC levels. Interference of the drug on the assay was assessed at 0.0, 0.1, 1.0, 100.0 μg/mL of tominersen in blank surrogate matrix and in surrogate matrix spiked at LLOQ and HQC levels. Interference of whole blood in the assay was assessed using increasing amounts of fully haemolysed whole blood from healthy volunteers (0, 0.1%, and 1.0% v/v) spiked in blank surrogate matrix and in surrogate matrix spiked at LLOQ and HQC levels.

Acceptance criteria for the absence of interference at a given concentration of interfering compound required ≥66.7% of the blank matrix aliquots (without reference standard) to have mean assay signals below LLOQ, and ≥66.7% of the spiked matrix samples to show accuracies within 70% and 130%.

The prozone effect (high-dose hook effect) describes a phenomenon observed in sandwich immunoassays in which the assay signal becomes saturated and falls in the presence of very high analyte concentrations (18). The prozone effect was assessed by spiking surrogate matrix with the highest attainable reference standard concentration above the ULOQ (12.5 μg/mL). The final amount of surrogate matrix was ≥95%. The spiked sample was serially diluted in surrogate matrix to bring at least one concentration within the assay working range (1 analysis in triplicates per dilution factor), i.e. sample analysed undiluted, and diluted 1 in 10, 1 in 100, 1 in 1,000, 1 in 10,000 and 1 in 100,000.

Stability of the reference standard in surrogate matrix was assessed at LQC and HQC levels. Samples were analysed in triplicate per concentration at the following conditions/time points: after 1 freeze/thaw cycle at –60 °C to –85 °C; after 2 and 4 hours at room temperature; after storage at –60 °C to –85 °C for approximately 1, 3, 6, and 12 months. Acceptance criteria required the accuracy of the mean concentration at each QC sample level to be within 70% and 130%; the precision of the mean concentration determined at each QC sample level needed to be ≤30% CV; and a maximum of one result per set was allowed to be rejected for analytical reasons.

Based on available assay development data, stability of the capture and detection antibodies mAb<mHTT>M-2B7-IgG-Bi lot BR02 and mAb<mHTT>M-MW1-IgG-Alexa647 lot BR08 was initially stated for 3 months of storage at 2–8 °C. The functional test consisting of the assessment of calibration curve performance was repeated after 3 months of storage at 2–8 °C. The stability of the capture and detection antibodies was considered acceptable if the signal-to-noise ratio at the LLOQ was ≥4; and the signal-to-noise ratio at the ULOQ was ≥1,000.

### Characterising assay specificity for mHTT

All recombinant HTT concentration responses were measured across a total of eight measurement plates. To compare responses across plates, the Q45 protein served as a reference signal that was measured on every plate. This allowed the normalisation of all plate responses according to the signal at its highest concentration. Parallelism was tested on two pre-dose CSF samples from patients with polyQ-repeat lengths of 41 and 42 (clinical study ISIS 443139-CS2), in addition to parallelism experiments conducted during assay validation.

Relative dose-responses were measured for recombinant HTT that varied in overall protein size, expression systems, vendors and polyQ-repeat numbers (**Table 5**). Assay signals were normalised using the HTT Q45 protein and protein concentrations expressed in fM.

To investigate suitability of the HTT Q46 reference standard for mHTTs with shorter polyQ-repeat lengths, parallelism was tested on two CSF samples prior to study drug injection, from patients with polyQ-repeat lengths of 41 and 42 (OLE of the Phase I/IIa study of tominersen). Study samples were serially diluted with surrogate matrix down to the LLOQ and below, with at least three different dilutions within the assay dynamic range and two below (e.g. MRD and additional 1:2, 1:3, 1:4, 1:6 and 1:8 dilutions). The recoveries were calculated based on concentration of the sample diluted at MRD.

### Statistics

All modelling analyses for the comparison of dose-responses curves across different recombinant HTT proteins were performed using RStudio v1.4.1717-3. Linear mixed effects models were fit using the lmer function from the lme4 package.

The null model with a random intercept for each unique recombinant protein was specified as:

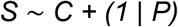

Where S is the measured assay signal, C is the nominal concentration and P is the unique recombinant proteins. This model was compared with the test model with random intercept and slope (nested model testing using ANOVA) which was specified as:

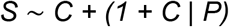

### Study approval

Samples were obtained from the OLE of the Phase I/IIa study of tominersen, which was approved by local ethics committees and conducted in accordance with the Declaration of Helsinki and the International Conference on Harmonization Guidelines for Good Clinical Practice.

The OLE study protocol was approved by the following ethics committees: NRES Committee London - West London and GTAC, London, UK; Ethik-Kommission der Medizinischen Fakultät der Universität Ulm, Germany; University of British Columbia Clinical Ethics Review Board, Canada.

Informed consent was provided by all patients prior to participation in the OLE study.

## Data Availability

The data that support the findings of this study are available from the corresponding author upon reasonable request.

## Author contributions

All authors contributed to the ongoing review and interpretation of the incoming data to inform the continued validation and finalisation of the assay.

SV, BM, and DJH designed the assay specificity experiments.

SV, MM, ES and LB designed and refined the assay strategy and validation experiments.

KS and AB conducted the assay development and validation experiments.

SV, BM and DJH analysed the data and wrote the manuscript.

DJH performed statistical evaluation of the assay specificity data.

NG supervised assay validation at ICON plc.

## Acknowledgments

Acknowledgements will be added at a later date.

## References

1. Roos RA. Huntington’s disease: a clinical review. Orphanet J Rare Dis. 2010;5(5):40.

2. Ross CA, Aylward EH, Wild EJ, et al. Huntington disease: natural history, biomarkers and prospects for therapeutics. Nat Rev Neurol. 2014;10(4):204–216.

3. Tabrizi SJ, Schobel S, Gantman EC, et al. A biological classification of Huntington’s disease: the Integrated Staging System. Lancet Neurol. 2022;21(7):632–644.

4. Aad G, Abbott B, Abdallah J, et al. Measurements of the Nuclear Modification Factor for Jets in Pb+Pb Collisions at radical(s)NN]=2.76 TeV with the ATLAS detector. Phys Rev Lett. 2015;114(7):072302.

5. Potter NT, Spector EB, Prior TW. Technical standards and guidelines for Huntington disease testing. Genet Med. 2004;6(1):61–65.

6. The American College of Medical Genetics/American Society of Human Genetics Huntington Disease Genetic Testing Working G. Laboratory guidelines for Huntington disease genetic testing. Am J Hum Genet. 1998;62:1243–1247.

7. Saudou F, Humbert S. The Biology of Huntingtin. Neuron. 2016;89(5):910–926.

8. Wild EJ, Boggio R, Langbehn D, et al. Quantification of mutant huntingtin protein in cerebrospinal fluid from Huntington’s disease patients. J Clin Invest. 2015;125(5):1979–1986.

9. Byrne LM, Rodrigues FB, Johnson EB, et al. Evaluation of mutant huntingtin and neurofilament proteins as potential markers in Huntington’s disease. Sci Transl Med. 2018;10:eaat7108.

10. Southwell AL, Smith SE, Davis TR, et al. Ultrasensitive measurement of huntingtin protein in cerebrospinal fluid demonstrates increase with Huntington disease stage and decrease following brain huntingtin suppression. Sci Rep. 2015;5:12166.

11. Fodale V, Boggio R, Daldin M, et al. Validation of Ultrasensitive Mutant Huntingtin Detection in Human Cerebrospinal Fluid by Single Molecule Counting Immunoassay. J Huntingtons Dis. 2017;6(4):349–361.

12. Tabrizi SJ, Leavitt BR, Landwehrmeyer GB, et al. Targeting huntingtin expression in patients with Huntington’s disease. N Engl J Med. 2019;380(24):2307–2316.

13. Ko J, Ou S, Patterson PH. New anti-huntingtin monoclonal antibodies: implications for huntingtin conformation and its binding proteins. Brain Res Bull. 2001;56(3-4):319–329.

14. World Health Organisation. UNDP/World Bank/WHO Special Programme for Research and Training in Tropical Diseases. Handbook: Good Laboratory Practice: Quality Practices for Regulated Non-clinical Research and Development. 2001.

15. Guilbault GG, Hjelm M. International Union of Pure and Applied Chemistry, Commission on Analytical Nomenclature and Clinical Chemistry Division. Nomenclature for automated and mechanised analysis, (recommendations 1989). Eur J Clin Chem Clin Biochem. 1991;29(9):577–585.

16. Tu J, Bennett P. Parallelism experiments to evaluate matrix effects, selectivity and sensitivity in ligand-binding assay method development: pros and cons. Bioanalysis. 2017;9(14):1107–1122.

17. Stevenson LF, Purushothama S. Parallelism: considerations for the development, validation and implementation of PK and biomarker ligand-binding assays. Bioanalysis. 2014;6(2):185–198.

18. Amarasiri Fernando S, Wilson GS. Studies of the ‘hook’ effect in the one-step sandwich immunoassay. J Immunol Methods. 1992;151(1-2):47–66.

