## Supplemental materials for "Quantifying mutant huntingtin protein in human cerebrospinal fluid to support the development of huntingtin-lowering therapies"

**Supplemental Figure 1. Concentration-assay response curves for HTT Q46 spiked in surrogate matrix or pooled human serum from healthy volunteers (remnant samples), processed and analysed on the same assay plate**.

aCSF, artificial CSF; CSF, cerebrospinal fluid; HTT, huntingtin protein.

**Supplemental Table 1. Antibody labelling outputs for 2B7 and MW1.**

| Labelling antibody and batch | Labelling date | Labelling ratio | Starting material   [mg] | Amount labelled antibody  [mg] | Yield   [%] | Purity   (SEC-HPLC) | Incorporation rate | Emission at 0.01 mg/mL |  | | | |
| --- | --- | --- | --- | --- | --- | --- | --- | --- | --- | --- | --- | --- |
| 2B7 BR01 | 28-Jan-2020 | NA | 1.0 | 0.48 | 48.0 | 99.7% | 0.7* | NA | x | x |  |  |
| 2B7 BR02 | 11-Mar-2020 | NA | 2.0 | 0.92 | 46.0 | 99.1% | 0.7* | NA |  |  | x | x |
| MW1 BR05 | 24-Feb-2020 | 1:3.5 | 0.32 | 0.27 | 84.4 | 98.4 | 2.8** | 272.1 |  |  |  |  |
| MW1 BR06 | 24-Feb-2020 | 1:5 | 0.32 | 0.27 | 84.4 | 98.3 | 3.9** | 343.6 |  |  |  |  |
| MW1 BR07 | 24-Feb-2020 | 1:7.5 | 0.32 | 0.27 | 84.4 | 98.1 | 5.5** | 376.3 | x |  | x |  |
| MW1 BR08 | 09-Mar-2020 | 1:8 | 2.0 | 1.72 | 86.0 | 98.4 | 5.9** | 411.2 |  | x |  | x |
| MW1 BR09 | 09-Jul-2020 | 1:8 | 2.0 | 1.66 | 79.5 | 98.3 | 5.9 | 412.5 |  |  |  |  |
| S/N |  |  |  |  |  |  |  |  | 5.2 | 7.1 | 5.2 | 6.2 |

* Label/IgG via ESI-MS. **** Label/IgG via absorbance.

S/N, response/background signal ratio at LLOQ of 1.63 pg/mL HTT Q46.

ESI-MS, electrospray ionization mass spectrometry; HTT, huntingtin protein; IgG, immunoglobulin G, LLOQ, lower limit of quantification; NA, not applicable; SEC-HPLC, size exclusion-high-performance liquid chromatography.

**Supplemental Table 2. Serial dilution of samples from patients with HD in surrogate matrix.**

| **Run** | **Sample dilution** | **Result before adjustment for dilution [pg/mL]** | **Dilution adjusted result [pg/mL]** | **Recovery based on undiluted sample [%]** | **Mean result [pg/mL]*** | **SD result [pg/mL]** | **%CV** |
| --- | --- | --- | --- | --- | --- | --- | --- |
| 1 | 1:1.1 | 9.84 | 10.8 | 100.0 | 7.22 | 0.45 | 6.3 |
|  | 1:2.2 | 3.41 | 7.50 | ***69.4*** |  |  |  |
|  | 1:3.3 | 2.03 | 6.70 | ***62.0*** |  |  |  |
|  | 1:4.4 | 1.70 | 7.47 | ***69.2*** |  |  |  |
| 2 | 1:1.1 | 7.46 | 8.21 | 100.0 | 8.80 | 1.02 | 12.4 |
|  | 1:2.2 | 3.98 | 8.76 | 106.7 |  |  |  |
|  | 1:3.3 | 3.01 | 9.93 | 121.0 |  |  |  |
|  | 1:4.4 | 1.88 | 8.28 | 100.9 |  |  |  |
|  | 1:6.6 | 1.08 | 7.11 | 86.6 |  |  |  |
|  | 1:11 | 1.03 | 11.3 | ***138.0*** |  |  |  |
| 3 | 1:1.1 | 5.04 | 5.54 | 100.0 | 5.60 | 0.55 | 9.8 |
|  | 1:2.2 | 2.34 | 5.14 | 92.8 |  |  |  |
|  | 1:3.3 | 1.89 | 6.24 | 112.6 |  |  |  |
|  | 1:4.4 | 1.36 | 5.96 | 107.6 |  |  |  |
|  | 1:6.6 | 0.730 | 4.81 | 86.8 |  |  |  |
|  | 1:8.8 | 0.673 | 5.93 | 107.0 |  |  |  |
| 4 | 1:1.1 | 6.49 | 7.13 | 100.0 | 7.15 | 0.62 | 8.7 |
|  | 1:2.2 | 3.21 | 7.05 | 98.9 |  |  |  |
|  | 1:3.3 | 2.49 | 8.23 | 115.4 |  |  |  |
|  | 1:4.4 | 1.68 | 7.39 | 103.6 |  |  |  |
|  | 1:6.6 | 0.988 | 6.52 | 91.4 |  |  |  |
|  | 1:8.8 | 0.750 | 6.60 | 92.6 |  |  |  |

* First outlier dilution excluded. Underlined values are below LLOQ of 1.63 pg/mL. ***Italic bold***: result outside ±30% recovery, not taken into account to calculate result mean and SD
CV, coefficient of variation; HD, Huntington’s disease; SD, standard deviation.

**Supplemental Table 3.** **Parallelism data for mHTT in CSF of patients with HD.** Serial dilution of samples from patients with low polyQ-repeat lengths (Q41 and Q42) in surrogate matrix.

| **Sample and polyQ-repeat length** | **Pre-dilution (excluding MRD of 1:2.2)** | **Sample pre-dilution (including on-plate dilution)** | **Results w/o dilution factor correction (pg/mL)** | **Dilution- adjusted result  (pg-eq/mL)** | **Recovery based on undiluted sample** | **Dilutions within the working range*** | |
| --- | --- | --- | --- | --- | --- | --- | --- |
| **1** | Undiluted | 1.1 | 3.49 | 3.83 | 100.0 | Mean | 4.00 |
|  | 1:2 | 2.2 | 1.77 | 3.90 | 101.8 | SD | 0.24 |
|  | 1:3 | 3.3 | 1.30 | 4.27 | 111.5 | %CV | 5.9 |
|  | 1:4 | 4.4 | BLQ | 4.80 | 125.3 |  |  |
|  | 1:6 | 6.6 | BLQ | 3.19 | 83.3 |  |  |
|  | 1:8 | 8.8 | BLQ | 2.56 | ***66.8*** |  |  |
| **2** | Undiluted | 1.1 | 3.42 | 3.76 | 100.0 | Mean | 3.97 |
|  | 1:2 | 2.2 | 1.77 | 3.89 | 103.5 | SD | 0.26 |
|  | 1:3 | 3.3 | 1.29 | 4.26 | 113.3 | %CV | 6.5 |
|  | 1:4 | 4.4 | BLQ | 4.82 | 128.2 |  |  |
|  | 1:6 | 6.6 | BLQ | 5.49 | ***146.0*** |  |  |
|  | 1:8 | 8.8 | BLQ | 6.08 | ***161.7*** |  |  |

***** Dilution level leading to concentrations slightly below LLOQ taken into consideration for precision calculation. **Italic bold**: result outside ±30% recovery. Underlined: result below LLOQ of 1.63 pg/mL, but result was taken into consideration for precision calculation.

BLQ, below LLOQ of 1.63 pg/mL; CSF, cerebrospinal fluid; CV, coefficient of variation; HD, Huntington’s disease; LLOQ, lower limit of quantification; mHTT, mutant huntingtin protein; MRD, minimum required dilution; NA, not applicable (result outside of acceptance criteria); polyQ, polyglutamine; SD, standard deviation; w/o, without.

**Supplemental Table 4. Microplate homogeneity of mHTT in surrogate matrix.** Accuracy of the LLOQ samples analysed.

|  | **Found mHTT concentration (pg/mL)** | | **Accuracy (%)** | |
| --- | --- | --- | --- | --- |
| **Sample ID** | **Roche** | **ICON** | **Roche** | **ICON** |
| 1 | 1.54^A^ | 1.63 | 94.5 | 99.6 |
| 2 | 1.74 | 1.38 | 106.7 | 84.1 |
| 3 | 1.68 | 1.19 | 103.1 | 72.5 |
| 4 | 1.73 | 1.55 | 106.1 | 94.3 |
| 5 | 1.84 | 1.37 | 112.9 | 83.7 |
| 6 | 1.70 | 1.79 | 104.3 | 109.2 |
| 7 | 1.61* | 1.69 | 98.8 | 103.0 |
| 8 | 1.35* | 1.57 | 82.8 | 95.5 |
| 9 | 1.42* | 1.77 | 87.1 | 107.8 |
| 10 | 1.93 | 1.68 | 118.4 | 102.5 |
| 11 | 1.72 | 1.53 | 105.5 | 93.1 |
| 12 | 1.58* | 1.77 | 96.9 | 107.8 |
| 13 | 1.82 | 1.47 | 111.7 | 89.9 |
| 14 | 1.78 | 1.53 | 109.2 | 93.4 |
| 15 | 1.56* | 1.55 | 95.7 | 94.7 |
| 16 | 1.99 | 1.42 | 122.1 | 86.3 |
| 17 | 1.50* | 1.79 | 92.0 | 109.2 |
| 18 | 1.85 | NA | 113.5 | NA |

* RO7234292 concentration extrapolated.

LLOQ, lower limit of quantification; mHTT, mutant huntingtin protein; NA, not available.

**Supplemental Table 5. Interference of wtHTT. Accuracy of specificity samples in surrogate matrix spiked with wtHTT (HTT-Q23 1-3144).**

| **Spiked HTT Q46 concentration (pg/mL)** | **Spiked HTT-Q23 1-3144 concentration (pg/mL)** | **Accuracy (%)** | |
| --- | --- | --- | --- |
|  |  | **Roche** | **ICON** |
| **Unspiked** | 200 | BLQ | BLQ |
|  |  | BLQ | BLQ |
|  |  | BLQ | BLQ |
|  | 20 | BLQ | BLQ |
|  |  | BLQ | BLQ |
|  |  | BLQ | BLQ |
|  | 0.00 | BLQ | BLQ |
|  |  | BLQ | BLQ |
|  |  | NA | BLQ |
| **1.63 (Roche)**  **1.64 (ICON)** | 200 | 129.4 | 105.9 |
|  |  | 118.4 | 136.3 |
|  |  | 141.1 | 121.2 |
|  | 20 | 124.5 | 90.1 |
|  |  | 133.7 | 115.6 |
|  |  | 118.4 | 97.2 |
|  | 0.00 | 100.6 | 98.9 |
|  |  | 106.7 | 104.5 |
|  |  | 117.8 | 103.2 |
| **400** | 200 | 100.3 | 95.5 |
|  |  | 92.3 | 54.4 |
|  |  | 91.3 | 81.4 |
|  | 20 | 95.0 | 78 |
|  |  | 88.8 | 91.3 |
|  |  | 95.0 | 84.4 |
|  | 0.00 | 98.5 | 108.9 |
|  |  | 98.0 | 81.9 |
|  |  | 99.8 | 108.5 |

Underlined values are out of acceptance criteria.

BLQ, below limit of quantification; HTT, huntingtin protein; NA, not available; wtHTT, wild-type HTT.

**Supplemental Table 6. Interference of drug. Accuracy of specificity samples in surrogate matrix spiked with tominersen.**

| **Spiked HTT Q46 concentration (pg/mL)** | **Spiked tominersen concentration (µg/mL)** | **Accuracy (%)** | | |
| --- | --- | --- | --- | --- |
|  |  | **Roche** | | **ICON** |
| **0** | 100 | BLQ | | BLQ |
|  |  | BLQ | | BLQ |
|  |  | BLQ | | BLQ |
|  | 1 | BLQ | | BLQ |
|  |  | BLQ | | BLQ |
|  |  | BLQ | | BLQ |
|  | 0.1 | BLQ | | BLQ |
|  |  | BLQ | | BLQ |
|  |  | BLQ | | BLQ |
| **1.63 (Roche)**  **1.64 (ICON)** | 100 | 103.1 | | 115.9 |
|  |  | 109.2 | | 117.8 |
|  |  | 112.3 | | 91.0 |
|  | 1 | 119.0 | | 185 |
|  |  | 96.3 | | 88.7 |
|  |  | 87.7 | | 75.2 |
|  | 0.1 | 120.2 | | 107.8 |
|  |  | 96.3 | | 105.1 |
|  |  | 95.1 | | 148.5 |
| **400** | 100 | 81.0 | 108.2 | |
|  |  | 88.0 | 82.0 | |
|  |  | 98.8 | 77.7 | |
|  | 1 | 91.5 | 58.7 | |
|  |  | 91.3 | 73.4 | |
|  |  | 97.0 | 73.8 | |
|  | 0.1 | 86.3 | 90.0 | |
|  |  | 100.3 | 79.4 | |
|  |  | 90.5 | 96.2 | |

Underlined values are out of acceptance criteria.

BLQ, below limit of quantification; HTT, huntingtin protein

**Supplemental Table 7. Interference of blood.** Accuracy of specificity samples in surrogate matrix spiked with haemolysed full blood.

|  |  | **Accuracy (%)** | |
| --- | --- | --- | --- |
| **Spiked HTT Q46 concentration (pg/mL)** | **Spiked full blood (v/v) (%)** | **Roche** | **ICON** |
| **0** | 1 | BLQ | Result in quantification range (10.8 pg/mL) |
|  |  | BLQ | Result in quantification range (9.40 pg/mL) |
|  |  | BLQ | Result in quantification range (6.86 pg/mL) |
|  | 0.1 | BLQ | BLQ |
|  |  | BLQ | BLQ |
|  |  | BLQ | BLQ |
|  | 0 | BLQ | BLQ |
|  |  | BLQ | BLQ |
|  |  | BLQ | BLQ |
| **1.63 (Roche)**  **1.64 (ICON)** | 1 | 138.0 | 482.3 |
|  |  | 116.6 | 636.4 |
|  |  | 126.4 | 482.1 |
|  | 0.1 | 106.1 | 115.4 |
|  |  | 120.2 | 116.6 |
|  |  | 110.4 | 108.7 |
|  | 0 | 95.7 | 142.6 |
|  |  | 90.2 | 97.7 |
|  |  | 128.2 | 116.3 |
| **400** | 1 | 147.5 | 98.9 |
|  |  | 100.3 | 94.9 |
|  |  | 121.3 | 95.1 |
|  | 0.1 | 105.0 | 95.4 |
|  |  | 110.8 | 116.7 |
|  |  | 94.3 | 121.9 |
|  | 0 | 94.0 | 103.8 |
|  |  | 99.0 | 70.7 |
|  |  | 123.3 | 91.4 |

Underlined values are out of acceptance criteria.

BLQ, below limit of quantification; HTT, huntingtin protein.

**Supplemental Table 8. Stability of reference standard in surrogate matrix, shock frozen and analysed after 4 hours storage at room temperature.**

| **Found mHTT concentration (pg/mL)** | | | | |
| --- | --- | --- | --- | --- |
|  | **LQC ( nominal concentration 4.50 pg/mL)** | | **HQC (nominal concentration 300.0 pg/mL)** | |
|  | **Roche** | **ICON** | **Roche** | **ICON** |
| Replicate 1 | 3.59 | 4.72 | 270 | 285 |
| Replicate 2 | 5.21 | 4.49 | 235 | 328 |
| Replicate 3 | 4.60 | 4.43 | 233 | 345 |
| **Mean conc. (pg/mL)** | 4.47 | 4.55 | 246 | 319 |
| **Accuracy (%)** | 99.3 | 101.1 | 82.0 | 106.5 |
| **Precision (%CV)** | 18.3 | 3.3 | 8.5 | 9.7 |

Conc., concentration; CV, coefficient of variation; HQC, high quality control; LQC, low quality control; mHTT, mutant huntingtin protein.

**Supplemental Table 9.** **Roche data: Stability of reference standard in surrogate matrix, shock frozen and analysed after 32 days of storage at –60 °C to** **–85 °C.**

| **Found mHTT concentration (pg/mL)** | | |
| --- | --- | --- |
|  | **LQC ( nominal concentration 4.50 pg/mL)** | **HQC (nominal concentration 300.0 pg/mL)** |
| Replicate 1 | 3.67 | 199 |
| Replicate 2 | 3.34 | 210 |
| Replicate 3 | 3.03 | 211 |
| **Mean conc. (pg/mL)** | 3.35 | 207 |
| **Accuracy (%)** | 74.4 | 69.0 |
| **Precision (%CV)** | 9.6 | 3.2 |

Conc., concentration; CV, coefficient of variation; HQC, high quality control; LQC, low quality control; mHTT, mutant huntingtin protein.

**Supplemental Table 10**. **ICON data: Stability of reference standard in surrogate matrix, shock frozen and analysed after 7, 28 and 85 days of storage at –70 °C.**

| **Found mHTT concentration (pg/mL)** | | |
| --- | --- | --- |
|  | **LQC ( nominal concentration 4.50 pg/mL)** | **HQC (nominal concentration 300.0 pg/mL)** |
| **Storage period: 7 days at –70 °C** | | |
| Replicate 1 | 4.52 | 227 |
| Replicate 2 | 3.84 | 267 |
| Replicate 3 | 4.39 | 231 |
| **Mean conc. (pg/mL)** | 4.25 | 242 |
| **Accuracy (%)** | 94.5 | 80.6 |
| **Precision (%CV)** | 8.5 | 9.2 |
| **Storage period: 28 days at –70 °C** | | |
| Replicate 1 | 6.04 | 298 |
| Replicate 2 | 4.44 | NA |
| Replicate 3 | 4.56 | 314 |
| **Mean conc. (pg/mL)** | 5.01 | 306 |
| **Accuracy (%)** | 111.4 | 102 |
| **Precision (%CV)** | 17.8 | 3.9 |
| **Storage period: 85 days at –70 °C** | | |
| Replicate 1 | 3.88 | 328 |
| Replicate 2 | 4.50 | 290 |
| Replicate 3 | 4.50 | 330 |
| **Mean conc. (pg/mL)** | 4.29 | 316 |
| **Accuracy (%)** | 95.4 | 105.3 |
| **Precision (%CV)** | 8.3 | 7.1 |

As one of the results may be rejected for analytical reasons, the results were accepted and there was no impact on the reliability of the data. Conc., concentration; CV, coefficient of variation; HQC, high quality control; LQC, low quality control; mHTT, mutant huntingtin protein; NA, not available – insufficient sample volume for analysis.
